# Multiplex PCR Assay to Detect High Risk Lineages of *Salmonella* Typhi and Paratyphi A

**DOI:** 10.1101/2021.09.14.21263553

**Authors:** Fahad A Khokhar, Derek DJ Pickard, Zoe A Dyson, Junaid Iqbal, Agila K Pragasam, Jobin Jacob John, Balaji Veeraraghavan, Farah N Qamar, Gordon Dougan, Hilary MacQueen, Sushila H Rigas, Mark A Holmes, Ankur Mutreja

## Abstract

Enteric fever infections remain a significant public health issue, with up to 20 million infections per year. Increasing rates of antibiotic resistant strains have rendered many first-line antibiotics potentially ineffective. Genotype 4.3.1 (H58) is the main circulating lineage of *S*. Typhi in many South Asian countries and is associated with high levels of antibiotic resistance. The emergence and spread of extensively drug resistant (XDR) typhoid strains has increased the need for a rapid molecular test to identify and track these high-risk lineages for surveillance and vaccine prioritisation. Current methods require samples to be cultured for several days, followed by DNA extraction and sequencing to determine the specific lineage. We designed and evaluated the performance of a new multiplex PCR assay, targeting *S*. Paratyphi A as well as the H58 and XDR lineages of *S*. Typhi on a collection of bacterial strains. Our assay was 100% specific for the identification of lineage specific *S*. Typhi and *S*. Paratyphi A, when tested with a mix of non-Typhi *Salmonella* and non-*Salmonella* strains. With additional testing on clinical and environmental samples, this assay will allow rapid lineage level detection of typhoid of clinical significance, at a significantly lower cost to whole-genome sequencing. To our knowledge, this is the first report of a SNP-based multiplex PCR assay for the detection of lineage specific serovars of *Salmonella* Typhi.

**Data Statement:** All supporting data, code and protocols have been provided within the article or through supplementary data files.

## INTRODUCTION

Enteric fever is caused by the bacteria *Salmonella enterica* serovars Typhi and Paratyphi A, B and C. It remains a significant health issue, which is estimated to cause up to 20 million infections and 161,000 deaths per year, predominantly in low and middle-income countries in Southeast Asia and Africa. *S*. Typhi and *S*. Paratyphi A are human restricted pathogens and are transmitted by the faecal-oral route often via contaminated water (1). Vaccination, access to clean water, and improved sanitation are effective means to prevent transmission of typhoid.

Cases of typhoid fever can be treated with former first-line antibiotics such as chloramphenicol, ampicillin or co-trimoxazole. However, the emergence of multi-drug resistant (MDR) *S*. Typhi in the mid-1970s and the recent emergence of strains with increased anti-microbial resistance (AMR) profiles to more antibiotics (2), has rendered these treatment options potentially ineffective in the near future. Many MDR *S*. Typhi strains possess self-transmissible incompatibility type (IncHI1) plasmids carrying a suite of antimicrobial resistance genes. Haplotype 58 (H58) or genotype 4.3.1 is the dominant *S*. Typhi lineage in many parts of Asia and Eastern Africa today and is associated with high levels of multidrug resistance and reduced susceptibility to fluoroquinolones (2).

Extensively drug-resistant (XDR) *S*. Typhi, first reported in Pakistan in 2016 (3), was found to be an MDR H58 strain that had acquired an IncY plasmid from an *E. coli* isolate harbouring both *bla*CTX-M-15 and *qnrS* resistance genes conferring resistance to ceftriaxone and third generation cephalosporins. This leaves azithromycin as the only viable oral treatment option for XDR *S*. Typhi. However, recent reports of azithromycin-resistant *S*. Typhi and *S*. Paratyphi A in Bangladesh (4) and Pakistan (5) have raised concerns about the potential of this mutation to evolve in XDR strains in the future, further limiting treatment options.

Current laboratory diagnosis of typhoid fever requires clinical samples to be sent to a centralised laboratory to be processed by bacterial culture and standard identification, followed by susceptibility, serological and/or advanced molecular tests such as real-time PCR and whole-genome sequencing (WGS). Blood culture remains the diagnostic technique of choice but takes several days for results and only identifies 45-70% of confirmed cases. It is also limited by the low numbers of *Salmonella* bacteria in samples ranging from <1-22 organisms/ml of blood (6, 7). Sampling from bone marrow has been shown to have greater sensitivity and specificity but that remains a highly invasive procedure. There have been several studies that utilise PCR assays to identify invasive *Salmonella* serovars (8-14) but rely on blood culture over 2-3 days followed by DNA extraction before PCR testing. More recently, studies have shown that the use of ox bile-containing media for the enrichment of bacteria directly from blood samples (15), or the selective removal of human DNA (16), combined with a PCR assay can reduce the turnaround time for diagnosis and increase the diagnostic sensitivity (17). Most importantly, however, is that none of the current routine diagnostic platforms highlighted here offer a resolution to discriminate between low and high-risk lineages in a single test.

In our study, we designed a simple, rapid, highly specific multiplex PCR assay for the detection of *S*. Typhi, *S*. Paratyphi A, *S*. Typhi H58 and *S*. Typhi XDR, based on single nucleotide polymorphisms (SNPs) specific for these lineages of clinical significance. Oncetested on clinical and environmental samples, this will be used to guide clinical and public health decision making.

## METHODS

### DNA extraction and bacterial strains

Bacterial isolates (Table 1) were cultured onto Luria-Bertani (LB) agar plates from frozen stock vials. A single bacterial colony was picked to subculture into 1 ml of LB broth and incubated overnight at 37°C in a shaking incubator. Genomic DNA was extracted from overnight cultures using the Wizard Genomic DNA Purification Kit (Promega, USA). Genomic DNA extracted from *S*. Typhi XDR strains was supplied by Aga Khan University, Karachi, Pakistan. All DNA strains were diluted in 10 mM Tris buffer and quantified using the Qubit 3 fluorometer with the dsDNA broad-range assay kit (Thermo Fisher Scientific, UK). An additional 76 DNA samples were obtained for screening using our PCR assay, that had been previously identified by Matrix-assisted laser desorption/ionization time-of-flight mass spectrometry (MALDI-TOF MS) as *S*. Typhi or *S*. Paratyphi A.

**Table 1.** Bacterial DNA strains used in testing. All *Salmonella* strains are serovars of *subspecies enterica* unless otherwise stated

| Bacterial strain | Isolate details |
| --- | --- |
| *S. Typhi* BRD948 | H10/15 Ty2 ($\Delta aroC aroD htrA$) |
| *S. Typhi* SGB87 | 98-0664 (H55) |
| *S. Typhi* | Quail strain |
| *S. Typhi* | lupe GEN0059 (4.1) |
| *S. Typhi* | 403 Ty (H59 3.1.2) |
| *S. Paratyphi* A | AKU_12601 |
| *S. Paratyphi* B | SPB7 |
| *S. Typhi* H58 | ERL12148 |
| *S. Typhi* H58 | ERL12960 |
| *S. Typhi* XDR 1 |  |
| *S. Typhi* XDR 2 |  |
| *S. Typhi* XDR 3 |  |
| *S. Typhi* XDR 4 |  |
| *S. Typhi* XDR 5 |  |
| *S. Enteritidis* | PT4 |
| *S. Hadar* | SC1 |
| *S. Infantis* | SC31 |
| *S. Cholearasuis* | SC-B67 |
| *S. enterica subsp. arizonae* | CDC 346-86 |
| *S. enterica subsp. diarizonae* | CDC 01-0005 |
| *S. Dublin* | TYT3627 |
| *S. Newport* | E2002001708 |
| *S. 1,4, (5),12:i* | CVM23701 |
| *S. Kentucky* | CVM29188 |
| *S. Heidelberg* | CVM30485 |
| *S. Javiana* | CVM35943 |
| *S. Saint Paul* | SARA29 |
| *S. Pullorum* | S449/87 |
| *S. Typhimurium* | D23580 |
| *S. Typhimurium* | 4/74 |
| *Escherichia coli* | BL21 |
| *Vibrio cholerae* | O1 El Tor |

### Primer design

Primers targeting highly conserved genes were designed for the detection of *S*. Typhi and *S*. Paratyphi A serovars (Table 2). Primers with additional mutations incorporated based on the mismatch amplification mutation assay (MAMA) PCR principle as previously described (18), were designed to selectively amplify a target sequence in the presence of a SNP of interest for H58 and XDR lineages (19). All primers were designed using Primer-BLAST (20), synthesised by Integrated DNA Technologies (IDT, USA) and were resuspended from their lyophilised form in 10 mM Tris buffer. Appropriate amplicon lengths were selected for each target with a minimum of 50 bp difference, in order to effectively separate and be visualised by gel electrophoresis.

**Table 2.**
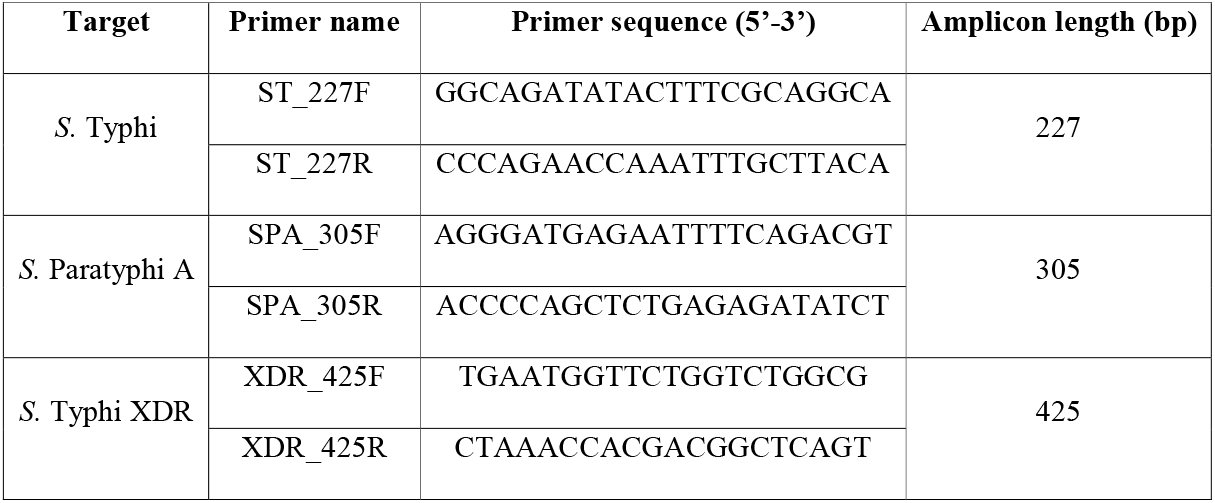
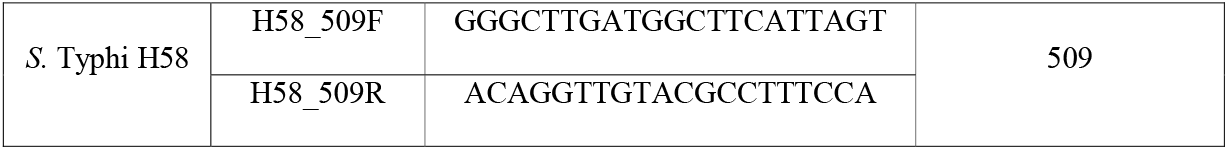
Primer sequences used in our multiplex assay with expected amplicon sizes generated for each target.

### PCR protocol

Each PCR reaction contained 5 ng of DNA, 12.5 μl of 2X PCR Master Mix (Thermo Fischer Scientific, UK), 8 or 10 μM each of the forward and reverse primers (Table 1), and nuclease free water to a final volume of 25 μl. All reactions were performed on a T100 thermal cycler (BioRad Laboratories Inc., USA) under the following cycling conditions; 95°C for 2 mins, followed by 30 cycles of 95°C for 30 sec, 60°C for 45 sec, 68°C for 1 min, and final extension at 68°C for 10 mins. The PCR products were run on 1.2% (w/v) agarose gels containing SYBR® Safe DNA Gel Stain (Invitrogen, USA) and visualised on a ChemiDoc MP imaging system (BioRad Laboratories Inc., USA).

## RESULTS

Our multiplex assay not only distinguished *S*. Paratyphi A from *S*. Typhi, but also identified low and high-risk lineages of *S*. Typhi that are of clinical relevance (Figure 1). Optimal conditions were initially determined for singleplex PCR, before adapting for the multiplex reaction. The *S*. Typhi and *S*. Paratyphi A primers were designed from specific regions in their reference genomes that are highly conserved across all strains, genes STY0307 and SSPA2308 respectively. The *S*. Typhi and *S*. Paratyphi A specific primers yielded 227 bp and 305 bp amplicon products, respectively.

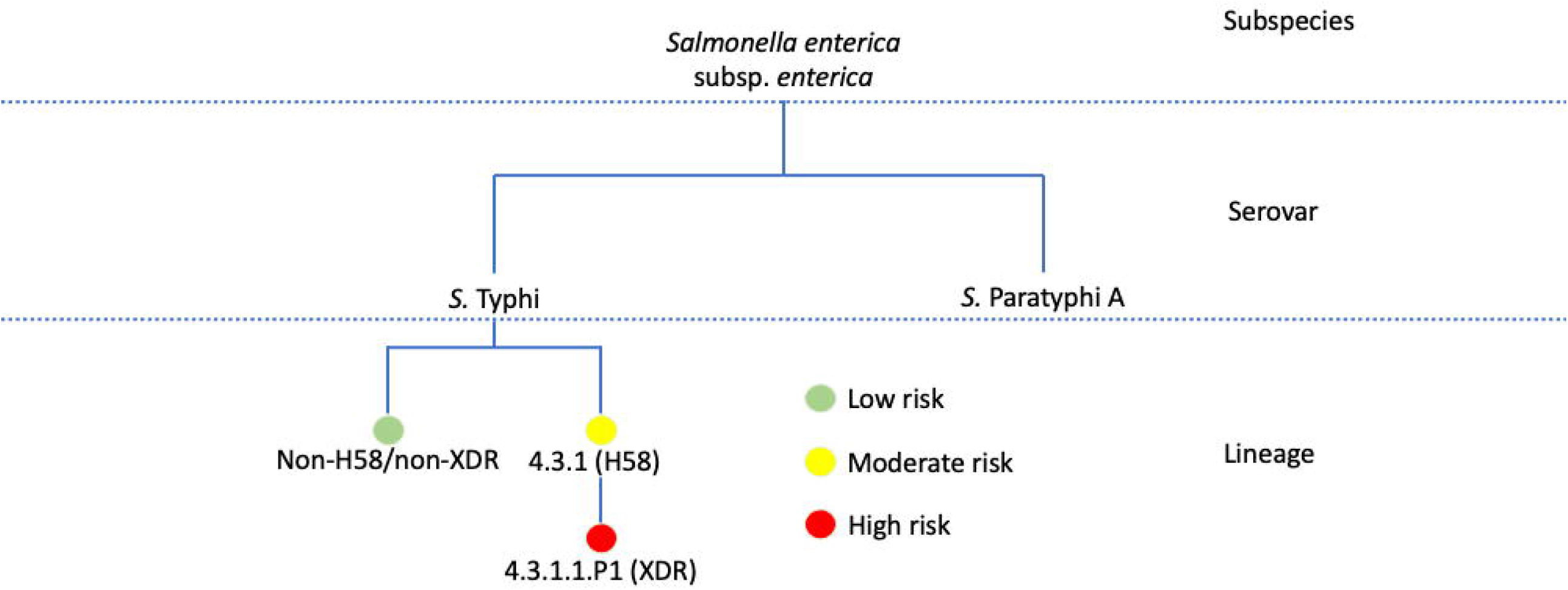

Through screening of our Typhi genome collection, we were able to determine SNPs which are specific to certain lineages, as detailed elsewhere (19, 21). These were chosen for use as relevant diagnostic markers. Our marker for the multidrug resistant lineage 4.3.1 (H58) is based on a C→T synonymous mutation (T349T) in the STY2513 gene at position 2348902 in the *S*. Typhi CT18 reference genome, which encodes for the anaerobic glycerol-3-phosphate dehydrogenase subunit A (glpA) gene. This mutation covers all H58 4.3.1 lineage and sub-lineage isolates, with our primers generating a 509 bp sized amplicon product. The diagnostic marker for the XDR lineage (genotype 4.3.1.1.P1) is based on a G→A (E13E) synonymous mutation in the STY0962 gene encoding anaerobic dimethyl sulfoxide reductase chain A precursor (dmsA), at position 955875 of the CT18 genome. Our XDR primers targeting this SNP generate an amplicon size of 425 bp. Figure 2 shows the results of the multiplex assay visualised by gel electrophoresis. Pairwise alignments to the CT18 reference sequence with our H58 and XDR SNPs are summarised in Figure 3 below.

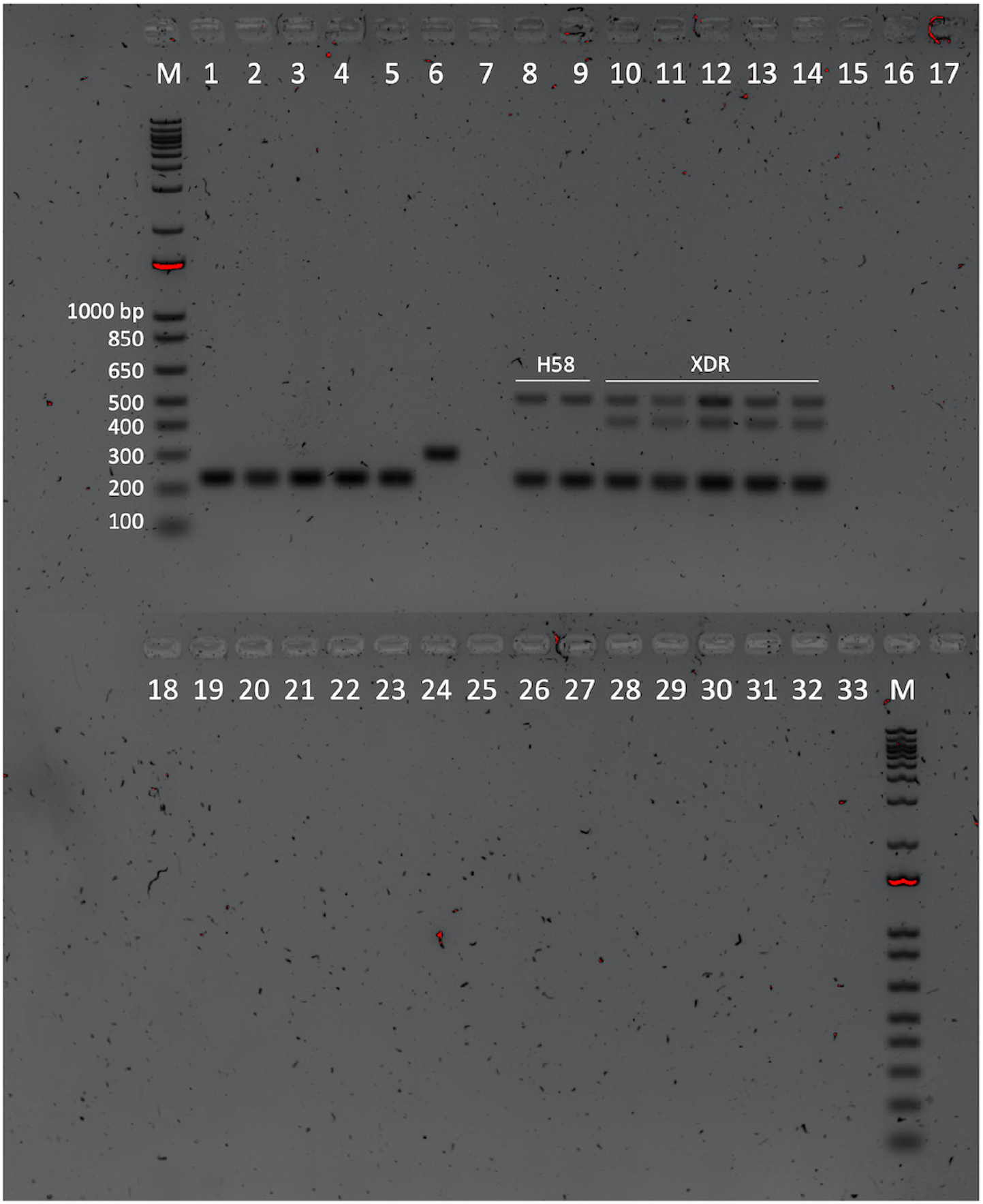

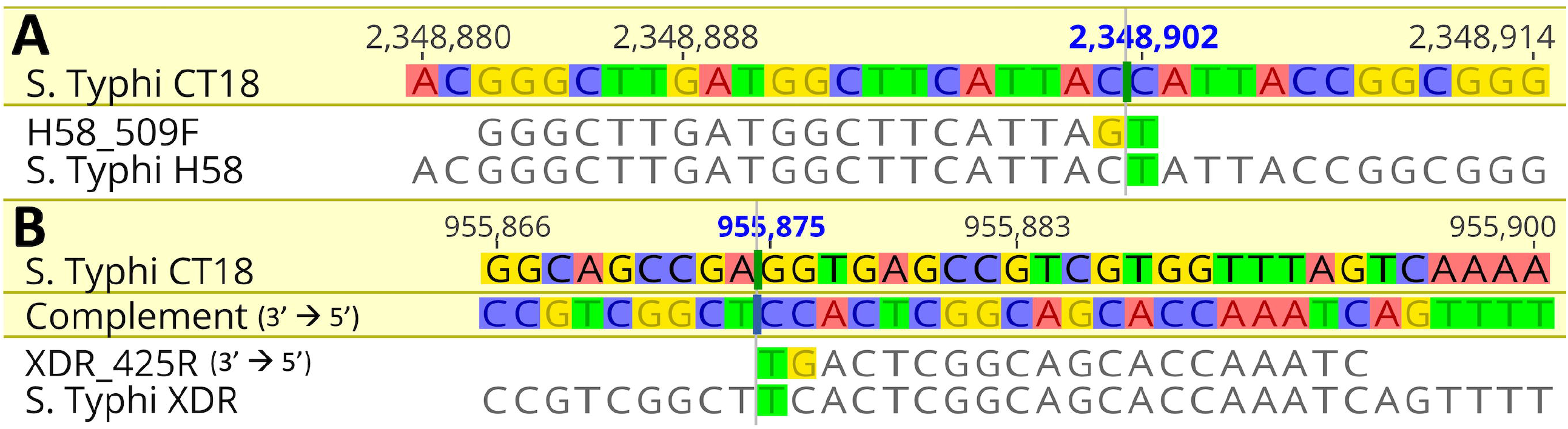

Our results showed that our initial primers designed for the H58 and XDR targets with just the one SNP incorporated were not specific for these lineages as they were binding to other *S*. Typhi strains (data not shown). Further adaptation was made by incorporating an additional SNP at the 3’ end of the primer which induced the specificity for the target sequence and strain and subsequently produced no false positive results (Figure 3). To determine the specificity of the assay, we tested the multiplex primers on a range of non-Typhi *Salmonella* and non-*Salmonella* pathogens, which showed no amplification.

From the additional DNA samples obtained for testing, 75/76 were originally identified as *S*. Typhi, with the remaining isolate identified as *S*. Paratyphi A. Using the same PCR conditions listed above, we tested our assay on these samples, with the single *S*. Paratyphi A sample confirmed by producing only a single 305 bp band as expected. Interestingly, 13/75 isolates that had previously been identified as *S*. Typhi, only produced the 305 bp *S*. Paratyphi A band in our assay. To investigate these discrepant results, the isolates were later whole-genome sequenced and confirmed as *S*. Paratyphi A. Of the remaining isolates, our PCR assay identified 54/76 as *S*. Typhi H58, 4/76 as *S*. Typhi non-H58/non-XDR, 1/76 as mixed *S*. Typhi and *S*. Paratyphi A, and 4/76 did not produce any bands (data not shown). None of these additional 76 DNA isolates were identified as *S*. Typhi XDR.

## DISCUSSION

Typhoid fever remains a major public health issue particularly in South and Southeast Asia. Early diagnosis is important for detecting cases in patients but also to discover sources of potential outbreaks. It is widely accepted that improved methods for the diagnosis and monitoring of the emergence and spread of *S*. Typhi would facilitate disease control and treatment. Yet there remain difficulties in obtaining sufficient samples for rapid molecular testing as direct blood samples are limited by the low abundance of *Salmonella* bacteria and other sampling methods remain extremely invasive and not widely available. Standard diagnostic techniques also lack the resolution to discriminate between different serovars of *S*. Typhi as well as drug-susceptible from drug-resistance strains. An ideal diagnostic for typhoid fever should be high-resolution, rapid, specific, and sensitive for the target organism.

The widespread dissemination of the H58 lineage in multiple countries, XDR lineage across Pakistan, and the emergence of azithromycin resistant isolates is of concern. The only method to identify and track the spread of such resistant strains is by whole genome sequencing. Based on our SNP-based diagnostic assay, there is potential to design and incorporate PCR primers as an additional target to monitor the spread of such strains at a significantly lower cost compared to complete genome sequencing.

Our multiplex PCR assay for the detection of *S*. Typhi and *S*. Paratyphi A enables the identification of the MDR H58 and XDR *S*. Typhi lineages. In many countries, *S*. Paratyphi A infections are increasingly common, so it was important that this assay could distinguish its presence in a population (22). Our single and multiplex PCR assays found no false positive reaction with non-Typhi serotypes or non-*Salmonella* pathogens, suggesting that the target genes are specific for our *Salmonella* serovar targets. From the additional DNA samples obtained, our assay was able to correctly identify *S*. Paratyphi A isolates that had previously been classified as *S*. Typhi by MALDI-TOF MS, which were subsequently confirmed as *S*. Paratyphi A by whole-genome sequencing. Although our assay was able to positively identify XDR *S*. Typhi from extracted DNA from five samples, we acknowledge there is a lack of additional XDR isolates in our collection. A further limitation is that our experiments were performed using DNA extracted from purified cultures and therefore contained few potential PCR inhibitors. Further optimisation of our PCR assay is planned for working directly on bacterial colonies in addition to sampling from environmental water and sewage samples. As direct assays are developed for clinical use in the future, our PCR assay should directly plug in to obtain the lineage level resolution.

We envision that our multiplex PCR assay will be widely used in routine laboratory diagnosis from DNA extracted after blood culture incubation, and eventually in combination with previously mentioned enrichment methods. Having a low cost, high-resolution, simple PCR test, as opposed to the current expensive whole-genome sequencing methods, allows our assay to be accessible to low- and middle-income countries where improved diagnostics for typhoid fever are most needed. The lineage level rapid detection of *S*. Typhi will make clinical decision making more efficient and help tackle AMR.

## Data Availability

All supporting data, code and protocols have been provided within the article or through supplementary data files.

## Author contributions

Conceptualisation and Funding Acquisition: A.M., M.A.H., G.D.; Investigation, Methodology and Visualisation: F.K. and D.J.J.P. Writing – Original Draft Preparation: F.K., D.J.J.P., M.A.H., A.M.; Writing – Review and Editing: all authors.

